# The spectrum of COVID-19-associated dermatologic manifestations: an international registry of 716 patients from 31 countries

**DOI:** 10.1101/2020.05.24.20111914

**Authors:** Esther E. Freeman, Devon E. McMahon, Jules B. Lipoff, Misha Rosenbach, Carrie Kovarik, Seemal R. Desai, Joanna Harp, Junko Takeshita, Lars E. French, Henry W. Lim, Bruce H. Thiers, George J. Hruza, Lindy P. Fox

**Author notes:** **Corresponding Author:** Esther Freeman, MD, PhD, Massachusetts General Hospital, 55 Fruit St, Boston, MA 02114.

## Abstract

**Question:** What are the cutaneous manifestations associated with COVID-19 and do they provide insight into the pathophysiology or prognosis?

**Findings:** In this international registry-based case series of 716 patients representing 31 countries, the most common dermatologic morphologies encountered in the 171 COVID-19 confirmed case included morbilliform, pernio-like, urticarial, macular erythema, vesicular, papulosquamous, and retiform purpura. Retiform purpura was seen exclusively in critically ill, hospitalized patients.

**Meaning:** COVID-19 is associated with a spectrum of skin findings in affected patients. These cutaneous manifestations may vary depending on the severity of COVID-19.

## Background

Coronavirus disease 2019 (COVID-19), caused by severe acute respiratory syndrome coronavirus-2 (SARS-CoV-2) has associated cutaneous manifestations.^1^ Case series have thus far documented lesions that are pernio-like,^2-6^ erythematous macules or papules,^5-7^ urticarial,^7,8^ morbilliform,^8^ varicelliform,^5-9^ and papulosquamous.^10^ Petechial eruptions,^8,11^ livedo reticularis-like rashes,^5,8^ purpuric lesions,^5^ and acro-ischemic lesions^12^ and retiform purpura^13^ have been reported less frequently. The timing of these varied eruptions in the disease course remains unclear, as do any potential associations between morphological subtypes with different COVID-19-associated syndromes, disease courses, and/or outcomes.

## Methods

In collaboration with the American Academy of Dermatology (AAD) and International League of Dermatologic Societies (ILDS), we established an international registry to collect cases of COVID-19 with cutaneous associations (www.aad.org/covidregistry); the registry is open to medical professionals only. No protected health information was collected, and all data was de-identified. The registry collected information on COVID-19 diagnosis type (suspected vs laboratory-confirmed), patient demographics, comorbidities, details regarding the patients’ new onset dermatologic condition, timing of dermatologic condition onset, information from skin biopsy reports when available, COVID-19 symptoms, and outcomes, including hospitalization, oxygen requirements, ventilator requirements, and deaths. All suspected or laboratory-confirmed COVID-19 cases, with new onset dermatologic findings, were eligible for inclusion in initial analysis. A sub-group analysis was performed restricted to lab-confirmed cases.

## Results

The registry collected 716 cases of new-onset dermatologic symptoms in patients with suspected or confirmed COVID-19 from April 8 to May 17, 2020. Cases came from 31 countries with most (89%) from the United States. Dermatologists entered most of the cases (54%). Median patient age was 30 years (IQR 19-49). Of the 171 patients in the registry with laboratory-confirmed COVID-19, the most common cutaneous morphologies, in order of frequency, were morbilliform, pernio-like, urticarial, macular erythema, vesicular, papulosquamous, and retiform purpura. In general, lesions tended to occur after (64%) or concurrently (15%) with other COVID-19 symptoms. Retiform purpura appeared in patients with the most severe disease course, presenting exclusively in very ill, hospitalized patients. Skin biopsies were reviewed for 15 patients.

## Conclusions and Relevance

This study highlights the array of cutaneous manifestations associated with COVID-19 diagnoses. Many morphologies were non-specific, while others may allow insight into potential immune or inflammatory pathways in COVID-19 pathophysiology. These cutaneous manifestations may vary depending on the severity of COVID-19, with retiform purpura exclusively reported in hospitalized patients.

## Data Availability

Data are housed through Partners healthcare Redcap and are not publicly available.

## Acknowledgements

We appreciate all the healthcare providers worldwide who entered cases in this registry. More detailed acknowledgements will appear in the published version of this paper.

## Ethics

The registry was reviewed by the Partners Healthcare (MGH) Institutional Review Board (IRB) and was determined to not meet the definition of Human Subjects Research.

